# Enhancing imagery rehearsal therapy for nightmares with targeted memory reactivation

**DOI:** 10.1101/2022.02.17.22270256

**Authors:** Sophie Schwartz, Alice Clerget, Lampros Perogamvros

## Abstract

Nightmare disorder (ND) is characterized by dreams with strong negative emotions occurring during rapid-eye movement (REM) sleep. ND is mainly treated by imagery rehearsal therapy (IRT), where the patients are asked to change the negative story line of their nightmare to a more positive one. We here used targeted memory reactivation (TMR) during REM sleep to strengthen IRT-related memories and accelerate remission of ND. Thirty-six patients with ND were asked to perform an initial IRT session and, while they generated a positive outcome of their nightmare, half of the patients were exposed to a sound (TMR group), while no such pairing took place for the other half (control group). During the next two weeks, all patients performed IRT every evening at home, and were exposed to the sound during REM sleep with a wireless headband, which automatically detected sleep stages. The frequency of nightmares per week at two weeks was used as the primary outcome measure. We found that the TMR group had less frequent nightmares and more positive dream emotions than the control group after two weeks of IRT, and a sustained decrease of nightmares after three months. By demonstrating the effectiveness of TMR during sleep to potentiate therapy, these results have clinical implications for the management of ND, with relevance to other psychiatric disorders too. Additionally, these findings show that TMR applied during REM sleep can modulate emotions in dreams.

## Introduction

Nightmares are characterized by the experience of strong negative emotions occurring usually during REM sleep. They involve images and thoughts of aggression, interpersonal conflict, and failure, and emotions like fear, anger and sadness ^1^. Nightmares may be idiopathic (without clinical signs of psychopathology) or associated with other disorders including post-traumatic stress disorder (PTSD). Posttraumatic nightmares are usually recurrent replications of the traumatic event, while idiopathic nightmares can have a variety of dream topics. Independently of their underlying origin, when nightmares are frequent and cause significant distress or impairment in social, occupational, or other important areas of functioning, they characterize the so-called nightmare disorder (ND), according to the international classifications of mental and sleep disorders (DSM-5, ICSD-3) ^2,3^. The average period prevalence rate of nightmares (at any given moment) at a clinically-significant level (>1 nightmare per week) is ∼4% of the general adult population across different epidemiological studies ^4-7^.

The pathophysiology and neural correlates of nightmares remain largely unknown. Two main etiological models of nightmares have been proposed ^8^: (i) increased hyperarousal and (ii) dysfunctional fear extinction. On the one hand, there is accumulating evidence that individuals with frequent nightmares have an increased number of arousals during sleep ^9,10^. Other studies have also found an increased sympathetic drive ^11^, elevated markers of emotional arousal ^12^, and increased alpha power during NREM and REM sleep ^13^ in patients with nightmares. Together, these observations suggest that ND shares some phenomenological and pathophysiological similarities with other disorders characterized by hyperarousal, such as insomnia disorder ^14^ and PTSD ^15^. However, other studies did not confirm the hyperousal model of ND ^16^.

On the other hand, it has been proposed that ND involves a dysfunction of fear extinction ^17^. As a reminder, fear extinction is defined as the gradual decrease of a conditioned response (e.g. avoidance), when a conditioned stimulus (CS) is presented without reinforcement from an unconditioned aversive stimulus (US) ^18^. During fear extinction, an inhibitory non-fearful (safety) memory that opposes the expression of the original fear memory is formed ^19^. Fear extinction is supported by a network that encompasses limbic regions (e.g., amygdala, medial prefrontal cortex, hippocampus, anterior cingulate cortex, insula), with the medial prefrontal cortex (mPFC) exerting an important inhibitory control on the amygdala ^20,21^. In the context of dreaming, it has been proposed that normal dreaming promotes fear extinction, by associating previous fearful memories (CS) with novel and dissociated contexts (in the dream itself) that do not predict the US ^17^. In a recent paper of our group ^22^, we showed that the more fear healthy participants had in their dreams, the less intense their fear response was while awake (i.e., decreased activity of amygdala and insula, paralleled by an increased activity of mPFC, when exposed to negative stimuli in the MRI scanner). This is consistent with extinction models according to which the mPFC exerts an inhibitory control over fear expression by reducing amygdala activity ^20^. Although this emotional function of dreams applies to negative but somewhat benign dreams, it may be disrupted in the case of distressing nightmares. Indeed, some recent studies showed that nightmare patients demonstrate a decreased mPFC activity during the viewing of negative pictures ^23^ and impaired frontal inhibitory functions ^24,25^. The aforementioned results support the fear extinction function of dreaming, and its failure in ND.

When untreated, ND can persist for decades ^26^. The only treatment with level A recommendation (i.e., based on strong evidence from randomized controlled trials and cohort studies) for ND is the Imagery Rehearsal Therapy (IRT) ^27^. IRT is a cognitive-behavioral technique that encompasses the following steps: recalling the nightmare, changing the negative story line, towards a more positive ending, and rehearsing the rewritten dream scenario during the day, which ultimately helps to reduce nightmares during sleep ^28^. This technique can be learned in one session ^29-31^, and practiced for 5-10 minutes per day while awake. A partial remission of nightmare frequency and severity has been observed after regular use of the technique for two to three weeks ^28,32^. Although IRT appears to be effective in the management of nightmares, approximately 30% of patients are unresponsive to this treatment ^28^. Therefore, new options to accelerate and enhance therapeutic outcome are needed.

Emerging evidence suggests that REM sleep favors the successful consolidation of extinction memory ^33-37^. Other studies showed that REM sleep helps to decrease the experienced affective tonus, thus leading to higher familiarity with emotionally negative stimuli (emotional depotentiation) ^38-40^ and to the consolidation of positive emotional memories ^41,42^. Developing experimental strategies aiming at leveraging these contributions of REM sleep could offer new therapeutic avenues for disorders with deficient emotion regulation (e.g., mood disorders, anxiety disorders, nightmares).

Targeted memory reactivation (TMR) is a technique used to modify memory formation through the application of cues during sleep ^43^. In a classic TMR protocol, a sensory (e.g., olfactory, auditory) cue is associated with a learning procedure during the day, and then administered during sleep. In that way, the replay of the associated memory and its corresponding neural representation in memory networks are artificially promoted, a procedure which will usually strengthens memory consolidation ^43^. TMR can improve declarative and procedural memory consolidation in humans ^44,45^, with the combined effect of sleep with TMR adding up to a memory enhancement of 35% as compared to wakefulness ^46^. While the beneficial effects of TMR in NREM for memory consolidation is undeniable ^47,48^, less is known about applying TMR in REM sleep. Some studies suggested that TMR in REM sleep enhanced memory ^49,50^, including the strengthening of associative emotional memory and memory generalization ^51^, and the reduction of negative valence of recently encountered stimuli ^38,52^. A recent study provided additional evidence that high-level cognitive processes take place during REM sleep as a response to external auditory stimuli ^53^. Importantly, sleep *alone* may not guarantee these emotional modulations; targeted potentiation of post-learning REM sleep seems important for such effects to be observed ^54^.

The current study included adults with ND and tested whether a TMR manipulation added to an IRT protocol accelerated remission of ND. More specifically, we investigated whether presenting, during REM sleep, a sound previously associated with a positive scenario of a nightmare (generated during an IRT session) decreased nightmare frequency (more than IRT alone). Specifically, we hypothesized that ND patients treated with IRT and who were exposed, during REM sleep and over 14 nights, to a sound that had previously been associated with the new positive dream scenario of IRT (TMR group), will have more reduced frequency of nightmares compared to participants with stimulation of the same, but non-associated, sound during REM sleep (control group). In the current TMR protocol, we associated a sound with a positive dream scenario, and therefore formulated an additional, secondary hypothesis according to which positive emotions should be reported more frequently after IRT in the dreams from the TMR group compared to the control group.

## Results

### Brief summary of Methods

Thirty-six patients with ND of at least moderate severity (>1 episode per week) according to ICSD-3 ^3^ were included. After an initial assessment and diagnosis of ND (pre-intervention), all participants filled in a sleep/ dream diary at home and wore an actimeter for 2 weeks (see design of the experiment illustrated in Figure 1). At the end of this period, all patients had a session of IRT. At the end of this session, half participants (TMR group) received a sound while imagining the new positive dream scenario, while the other half did not receive the sound (control group). During the following 2 weeks, all patients received the sound during REM sleep with a wearable headband EEG device (Dreem^®^) that automatically scored sleep stages and delivered the sound every 10 seconds during REM sleep specifically. During these 2 weeks, all participants performed rehearsal of the positive dream scenario for 5 minutes and filled in a dream diary. At the end of this period, they had a second assessment session (post-intervention) about the evolution of their ND. Finally, a third assessment session (3-month follow-up) took place 3 months later. We asked the participants to retrospectively report the number of nightmares per week for the last 2 weeks at pre-, post- and follow-up (primary outcome measure). Second outcome measures included the proportions of joy and fear in dreams, sleep efficiency (SE), as measured by actigraphy (SE_Acti_) and the Dreem EEG headband (SE_Dreem_), as well as scores in the Nightmare Distress Questionnaire (NDQ), the Pittsburgh Sleep Quality Index (PSQI) and the Beck Depression Inventory (BDI-II) questionnaire.

**Figure 1.**
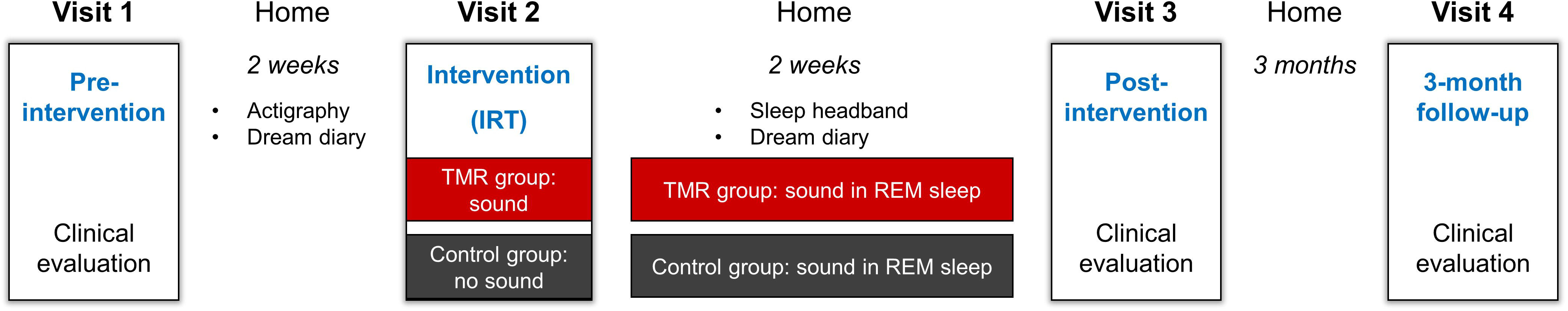
Design of the Experiment. Patients with ND had a first clinical assessment of nightmare intensity and frequency with standardized questionnaires at time Visit 1 (pre-intervention), after which they filled in a sleep and dream diary at home and wore an actigraph for two weeks. At the end of this period, they had an Imagery Rehearsal Therapy (IRT) session (Visit 2). At the end of this session, patients imagined the new positive ending of their nightmare either in the presence of a contextual sound (TMR group) or not (control group). For the following two weeks, all patients (TMR group, control group) were presented the sound during REM sleep at home with a headband device, while filling in a dream diary every morning. Nightmare intensity and frequency was measured again after two weeks of TMR at home (Visit 3, post-intervention), and 3 months later (Visit 4, follow-up). Dream diaries were collected continuously between Visit 1 and Visit 3.

Recruitment took place from May 2020 to October 2021, with final follow-up data collected in January 2022. Supplementary Figure 1 provides a flow diagram of the study participants.

### Baseline demographic, sleep and dream characteristics

At pre-intervention, no significant baseline differences were found between the groups for demographic and clinical characteristics, including primary and secondary outcome variables (Table 1).

**Table 1.** Baseline demographic, sleep and dream characteristics.

|  | TMR group<br>(N=18) | Control group<br>(N=18) | df | t | p-value | d |
| --- | --- | --- | --- | --- | --- | --- |
| Age (years) | 26.11 ± 3.67 | 24.88 ± 3.86 | 34 | 1.16 | 0.25 | 0.32 |
| Female sex | 13 | 14 | 1 | 0.14 | 0.70 | 0.06 |
| Nightmares/week<sub>pre</sub> | 2.94 ± 1.73 | 2.58 ± 1.19 | 34 | 0.38 | 0.72 | 0.24 |
| BDI-II<sub>pre</sub> | 14.55 ± 7.72 | 12.83 ± 8.80 | 34 | 0.82 | 0.42 | 0.20 |
| NDQ<sub>pre</sub> | 27.61 ± 7.53 | 26.55 ± 7.65 | 34 | 0.41 | 0.67 | 0.13 |
| PSQI<sub>pre</sub> | 7.77 ± 2.86 | 7.55 ± 3.22 | 34 | 0.22 | 0.82 | 0.07 |
| SE<sub>Acti</sub> (%) | 84.96 ± 6.62 | 87.68 ± 7.85 | 34 | -1.12 | 0.27 | 0.39 |
| JOY<sub>pre</sub> | 0.12 ± 0.24 | 0.17 ± 0.18 | 34 | -0.64 | 0.52 | 0.23 |
| FEAR<sub>pre</sub> | 0.57 ± 0.33 | 0.62 ± 0.24 | 34 | -0.55 | 0.58 | 0.17 |
| TST<sub>Acti</sub> | 426.30 ± 41.25 | 435.26 ± 73.2 | 34 | -0.45 | 0.65 | 0.15 |

*Means, standard deviations and comparison between the control and TMR group, for age, female sex and outcome variables at pre-intervention. Values are reported as mean ± SD. For each variable, the degrees of freedom, Student's t value ( $X^2$ for female sex), the p-value and Cohen's d values (Cramér's V for female sex) are reported.*
*Pre: pre-intervention, BDI-II: Beck Depression Inventory, NDQ: Nightmare Distress Questionnaire, PSQI: Pittsburgh Sleep Quality Index, JOY<sub>pre</sub> / FEAR<sub>pre</sub>: proportion of dreams with joy and fear, respectively, among all remembered dreams during two weeks before IRT, TST<sub>Acti</sub>: Amount of sleep from lights out (recording starts) to lights on (recording ends), SE<sub>Acti</sub>: sleep efficiency = $TST_{Acti} / TBT_{Acti}$ (TBT<sub>Acti</sub>: duration between lights on and lights off).*

### Sound stimulation at home

No significant differences between the groups were observed for the average number of stimulations, average sound intensity, sleep efficiency (SE_Dreem_), sleep macrostructure and REM arousal index (number of arousals in stage REM × 60/REM duration) during the two weeks of IRT and stimulations, as measured by the sleep headband (Table 2).

**Table 2.**
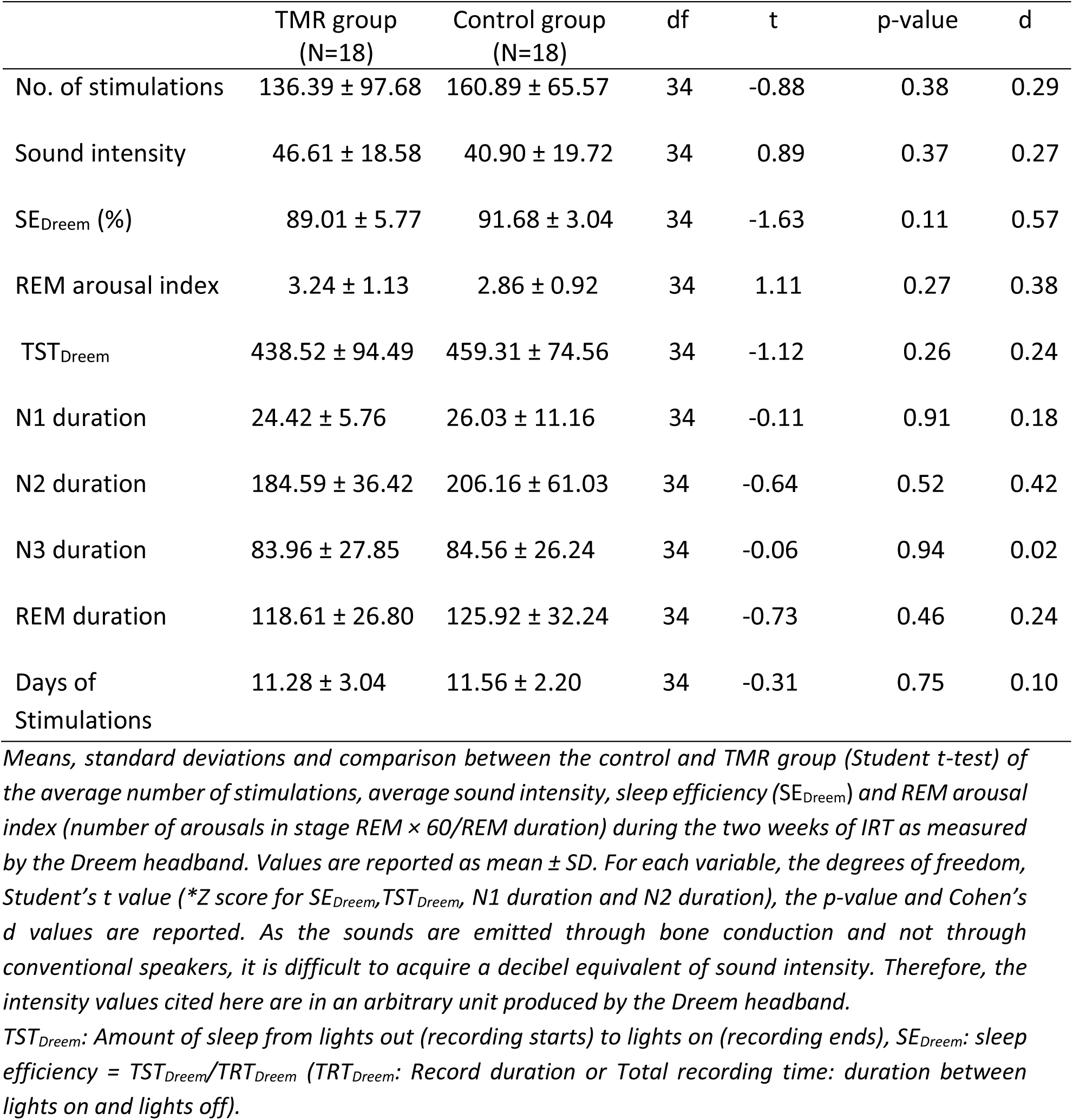
Comparison between groups during auditory stimulations.

|  | TMR group<br>(N=18) | Control group<br>(N=18) | df | t | p-value | d |
| --- | --- | --- | --- | --- | --- | --- |
| No. of stimulations | 136.39 ± 97.68 | 160.89 ± 65.57 | 34 | -0.88 | 0.38 | 0.29 |
| Sound intensity | 46.61 ± 18.58 | 40.90 ± 19.72 | 34 | 0.89 | 0.37 | 0.27 |
| SE<sub>Dreem</sub> (%) | 89.01 ± 5.77 | 91.68 ± 3.04 | 34 | -1.63 | 0.11 | 0.57 |
| REM arousal index | 3.24 ± 1.13 | 2.86 ± 0.92 | 34 | 1.11 | 0.27 | 0.38 |
| TST<sub>Dreem</sub> | 438.52 ± 94.49 | 459.31 ± 74.56 | 34 | -1.12 | 0.26 | 0.24 |
| N1 duration | 24.42 ± 5.76 | 26.03 ± 11.16 | 34 | -0.11 | 0.91 | 0.18 |
| N2 duration | 184.59 ± 36.42 | 206.16 ± 61.03 | 34 | -0.64 | 0.52 | 0.42 |
| N3 duration | 83.96 ± 27.85 | 84.56 ± 26.24 | 34 | -0.06 | 0.94 | 0.02 |
| REM duration | 118.61 ± 26.80 | 125.92 ± 32.24 | 34 | -0.73 | 0.46 | 0.24 |
| Days of<br>Stimulations | 11.28 ± 3.04 | 11.56 ± 2.20 | 34 | -0.31 | 0.75 | 0.10 |

*Means, standard deviations and comparison between the control and TMR group (Student t-test) of the average number of stimulations, average sound intensity, sleep efficiency (SE<sub>Dreem</sub>) and REM arousal index (number of arousals in stage REM × 60/REM duration) during the two weeks of IRT as measured by the Dreem headband. Values are reported as mean ± SD. For each variable, the degrees of freedom, Student's t value (\*Z score for SE<sub>Dreem</sub>, TST<sub>Dreem</sub>, N1 duration and N2 duration), the p-value and Cohen's d values are reported. As the sounds are emitted through bone conduction and not through conventional speakers, it is difficult to acquire a decibel equivalent of sound intensity. Therefore, the intensity values cited here are in an arbitrary unit produced by the Dreem headband.*
*TST<sub>Dreem</sub>: Amount of sleep from lights out (recording starts) to lights on (recording ends), SE<sub>Dreem</sub>: sleep efficiency = TST<sub>Dreem</sub>/TRT<sub>Dreem</sub> (TRT<sub>Dreem</sub>: Record duration or Total recording time: duration between lights on and lights off).*

### Clinical outcome variables

#### Primary outcome variable

There was a significant Time*Group interaction (p=.003, F=6.50) and a significant effect of Time (p=<.001, F=60.11) for the number of nightmares/week (Figure 2A). A pairwise t-test indicated that the TMR group had significantly lower nightmares/week than the control group at post-intervention (*p* = .021, t=-2.34, Cohen’s d= 0.79) and the 3-month follow-up (*p* = .007, t=-2.76, Cohen’s d= 1.03) (Table 3).

**Figure 2.**
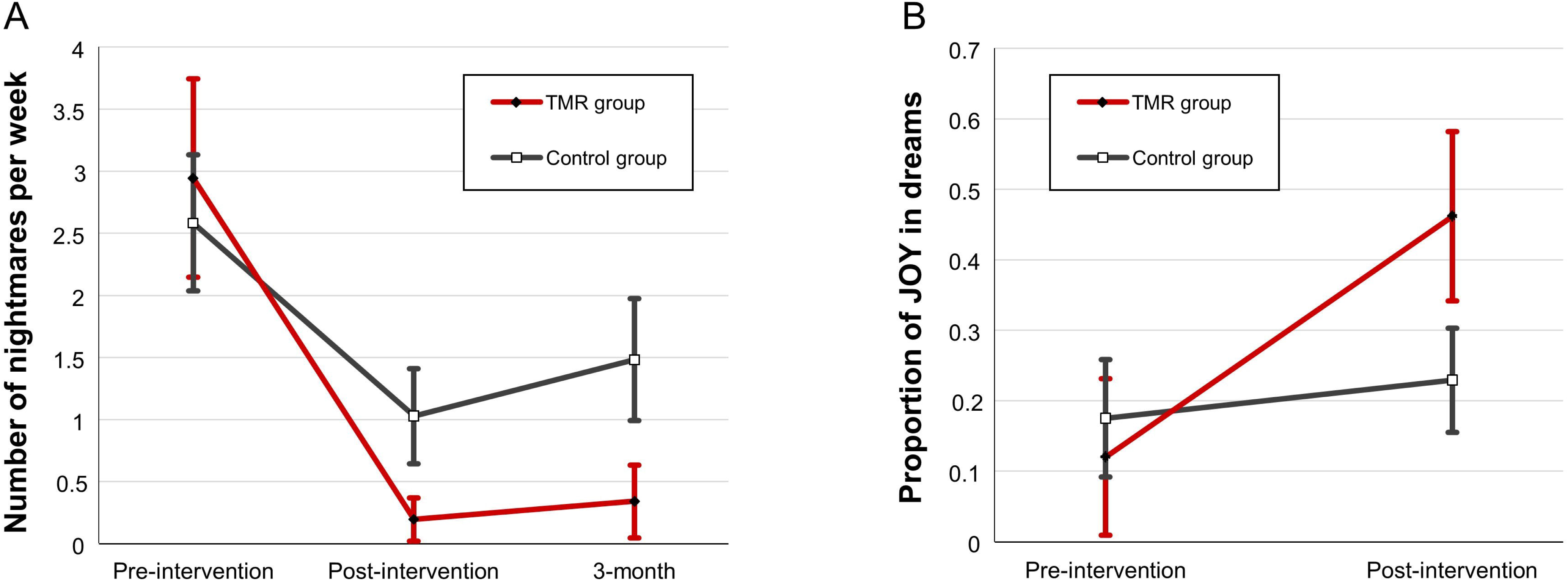
Treatment effects on (A) the number of nightmares per week and (B) proportion of joy in dreams. Data for the TMR and the control groups at baseline (pre-), two weeks after IRT (post-) and at the 3-month follow-up (3-month). Error bars represent 95% CI.

**Table 3.** Comparison between groups at post-intervention and 3-month follow-up.

|  | TMR group<br>(N=18) | Control group<br>(N=18) | df | t | p-value | d |
| --- | --- | --- | --- | --- | --- | --- |
| Nightmares/week<sub>post</sub> | 0.19 ±0.38 | 1.02 ±0.83 | 77.1 | -2.34 | 0.021* | 0.79 |
| Nightmares/week<sub>3-m</sub> | 0.33 ±0.56 | 1.48 ±0.94 | 86.3 | -2.76 | 0.007* | 1.03 |
| JOY<sub>post</sub> | 0.46±0.26 | 0.22±0.16 | 53.6 | 3.26 | 0.002* | 1.15 |

*Values are reported as mean ± SD; post: post-intervention; 3-m: 3-month follow-up; , JOY<sub>post</sub>: proportion of dreams with joy among all remembered dreams during the two weeks of IRT. Pairwise comparisons with Holm-Sidak correction for the outcome variables with a significant Time\*Group interaction. \*p<.05*

#### Secondary outcome variables

There was a significant Time*Group interaction (p=<001, F=16.95) and a significant effect of Time (p=<.001, F=32.03) for the emotion of joy in dreams, but no effect of group (p=.16, F=2.05) (Figure 2B). Pairwise t-tests indicated that the TMR group had significantly higher proportion of joy than the control group at post-intervention (*p* = .002, t=3.26, Cohen’s d= 1.15) (Table 3), and that only the TMR group had higher joy in dreams at post-intervention compared to pre-intervention (p=<.001, t=-6.91), whereas the control group had no such time effect (p=.28, t=-1.09). There was a significant effect of Time for the proportion of fear (p=<.001, F=19.69), but no Group effect (p=.69, F=0.15) or Time*Group interaction (p=.63, F=0.23) (Supplementary Figure 2). All other exploratory variables also displayed an effect of Time (NDQ: p=<.001, F=20.81; PSQI: p=<.001, F=12.60; BDI-II: p=.003, F=6.34; SE: p=.002, F=10.53), and no Group effect (NDQ: p=.85, F=0.03; PSQI: p=.99, F=0.01; BDI-II: p=.36, F=0.83; SE: p=.10, F=2.81) or Time*Group interaction (NDQ: p=.44, F=0.81; PSQI: p=.90, F=0.10; BDI-II: p=.92, F=0.08; SE: p=.98, F=0.0003) (Supplementary Figure 2).

In order to test for a relationship between the emotions of fear and joy in dreams, we performed correlational analyses of these emotions at post-intervention. The TMR group showed a significant negative correlation (tau=-.47, p=.007), while this correlation was not significant for the control group (tau=-.12, p=.49). After transformation of tau values to Pearson r-values ^55^, a Fisher r-to-z transformation test showed that the correlation coefficients were significantly different between the groups (p=0.046, z=-1.68).

A supplementary analysis of the content of the 531 dream reports from the dream diary using the Linguistic Inquiry and Word Count (LIWC) software (https://www.liwc.app/ ^56^) confirmed that the TMR group had significantly higher positive emotions in dreams than the control group at post-intervention (p = .04, t=2.04), while there was no significant difference between groups at pre-intervention (p = .95, t=0.05; significant Time*Group interaction, p=.01, F=6.17; see Supplementary Material S3). No such effect was observed for negative emotions.

Finally, we showed that the increase in positive emotions in the TMR group was unlikely attributable to a veridical replay of the IRT-related scenario or elements. Indeed, in order to compare the IRT script to subsequent dream scripts (n=220), we used latent semantic analysis (LSA), which extracts meaningful similarities between texts ^57^, and found very low similarity scores (see Supplementary Material S4).

#### Effect of stimulations on REM sleep

When comparing the first two nights of stimulations with the last two nights for the REM arousal index, there was no significant Time*Group interaction (p=.851, F=.036), no effect of Time (p=.929, F=008) or Group (p=.221, F=1.56), meaning that stimulation (associated or not) did not alter REM sleep structure.

## Discussion

We here demonstrate a clinical reduction of nightmare frequency in a population of nightmare sufferers under combined IRT and TMR compared to patients under IRT alone. Moreover, we report an increase in dreams containing positive emotions (i.e., joy) in the TMR group only. Together, these findings suggest that TMR during REM sleep may improve IRT outcome by favoring the activation of emotionally positive dreams, while significantly decreasing nightmares.

We first show that IRT was effective for patients from both the TMR and control groups, with a significant effect of time across all variables (nightmare frequency, joy and fear in dreams, distress, mood scores, sleep quality). These results confirm previous reports showing that IRT reduces nightmare frequency and distress ^58,59^, negative emotions in dreams ^60^, while improving sleep quality ^58,61^, insomnia severity ^62^ and mood scores ^58,63^. However, although IRT is the established treatment of nightmares since more than 30 years now ^30^, the lack of a control group (without IRT) in the current study design cannot exclude that the clinical improvements observed in both patients groups were influenced by other factors (e.g., patient expectations).

Critically, the frequency of weekly nightmares was further reduced in the TMR compared to the control group, thus supporting our hypothesis that TMR is an efficient method to potentiate IRT. This difference displayed a medium to large effect size and was sustainable at the 3-month follow-up. The role of TMR in the consolidation of positive associative memories during REM sleep, reflected by the significant increase of positive emotions (i.e., joy) in dreams of the TMR group, may provide a plausible mechanistic explanation for the therapeutic benefit of TMR in this study. As TMR did not further reduce nightmare distress or fear in dreams, the contributions of IRT and TMR to alleviating nightmares in this specific study may be partly dissociated, with IRT mostly inhibiting fear in dreams (e.g., fear extinction, rendering threatening stimuli more neutral), while TMR boosting emotionally positive dreams, as we further discuss below.

On the one hand, some authors also proposed that IRT may increase the feeling of mastery over dream content ^60,64^. Indeed, the increase of control over the content of nightmares through mental imagery has been described as an important component of the IRT program ^28^. On the other hand, IRT may also directly affect emotion regulation systems (e.g., extinction learning) ^65^, a mechanism that is compatible with theoretical models of dreams and nightmares ^17,66^. Indeed, dreams serve a fear extinction function by exposing the individual to feared stimuli (conditioned stimuli-CS) in a safe and virtual environment that does not predict the unconditioned stimulus-US ^17^. This procedure (CS-noUS associations and violation of expectancy) is related to extinction and inhibitory learning ^18^. Therefore, rescripting a nightmare with a positive scenario, as in IRT, would represent a mental rehearsal of CS–noUS associations ^18^, with the concurrent presence of a feared stimulus and a tolerable outcome. IRT would then share similar mechanisms with exposure therapy ^32,63^ by enhancing extinction and rendering threatening stimuli more neutral.

Importantly, IRT seems to be related to the application of an emotional skill learned during wakefulness (e.g., reappraisal) and which is then generalized in other situations during the following dreams. This is supported by the clinical observation that the individual under IRT is rarely dreaming of the new scenario ^65^, which we confirmed by applying the latent semantic analysis (LSA) on the dreams of our patients (Supplementary Material S4). The results of this analysis suggest that direct incorporations of the IRT scenario in the dreams that the patients reported in the morning were exceptional. The changes in dream emotionality and the related differences between the groups may thus not be attributable to a veridical replay of the IRT-related scenario or elements.

In a similar way, it has been shown that TMR during sleep promotes memory generalization^51^ and the consolidation of categorically-related memories, but not necessarily the consolidation of one specific memory trace ^67^. Interestingly, IRT with TMR during REM sleep in our protocol decreased the frequency of nightmares and increased positive emotions in dreams, but did not further decrease fear in dreams compared to controls with IRT only. This suggests that positive dreams progressively took over nightmares as the most prevalent type of dreams in the patients of the TMR group, which is supported by both the subjective scoring of emotions in dreams by the patients, the correlational analysis between fear and joy in dreams at post-intervention, and an automated (i.e., objective) lexico-statistical analysis of dreams (LIWC) (Supplementary Material S3). Recent studies demonstrated that TMR during REM sleep helps consolidate emotional associative memories and generalization ^51^, while it also increases positive valence of initially negative stimuli ^38,52^. Associating a positive scenario (during IRT) with a sound, as we did here, and replaying this association during REM sleep with TMR over several consecutive nights would then promote the long-term consolidation of positive memory traces and memory generalization. This interpretation is also supported by the sustained effect observed after 3 months. We can therefore assume that a TMR intervention of 2 weeks is effective for long-term benefits on nightmare reduction. Of course, future studies may further investigate whether other intervention periods (shorter or longer) may result in varying long-term effects of the treatment.

Concurring with scarce previous research ^41,42^, this study provides experimental evidence for a role of REM in the consolidation of positive associative memories. The underlying mechanisms of IRT and TMR may have converging (possibly additive) effects on the emotionality of the dream experience, and not strictly to the specific changes in dream emotionality implied by each of these mechanisms. For instance, reducing fear in dreams (as caused by a fear extinction process) could foster the expression of positive emotions in dreams. Besides, our measurements were taken over the 2 weeks of intervention and it may be that IRT alone also increases positive emotions in dreams later on during therapy ^60^. In any case, our results suggest that TMR acts as a booster in this procedure.

There are certain limitations in this study. First, the current design lacked a group of nightmare patients without any stimulations. As mentioned above, the goal of the study was not to demonstrate the efficacy of the IRT itself, which is already well established for ND, but to test for any additive effect of TMR. Yet, because we did not acquire polysomnographic measures before stimulations, the current design does not allow assessing other possible unspecific effects of the sound stimulation on sleep. Moreover, to control for any unspecific effect of the sound played during IRT (e.g., S1, in the TMR group), future studies may use a different sound (e.g., S2) played during IRT in an additional control group, while the same sound (S1) would be played during sleep of both groups. This sound (S2) in the control group would need to be similar in terms of valence and intensity to the sound (S1) in the TMR group, but sufficiently different to minimize generalization between the two sounds in the control group. Furthermore, some prior work has suggested that the mere expectation of future reward (for good memory performance) may boost sleep-related memory consolidation processes ^68^. Future studies could therefore test whether prior explicit knowledge (e.g., via instruction) about the link between sound during IRT and during sleep may further boost the effect of TMR during sleep, and could thus be added to TMR-based therapies. Finally, another important aspect concerns the dropout rate at the 3-month follow-up evaluation (22%), which is at the superior limits of general acceptability for long-term randomized trials (20%) ^69^, although such a rate is expected in this specific group of clinical population ^58^. Please note, however, that this rate is lower than most studies using psychological treatments for nightmares, where the dropout rates range from 23% to 52% ^70^.

The present study establishes that associating TMR to IRT reduces nightmares frequency and fosters positive emotion in dreams. Such a TMR manipulation could accelerate a process whereby dreams regain their functional role of fear extinction. Thus, we propose that TMR in REM sleep could be used as a new *‘sleep therapy’* in other psychiatric disorders with deficient extinction/emotion regulation as well (e.g., anxiety disorders, PTSD, mood disorders, insomnia disorder) ^71,72^. By deploying and popularizing easy-to-use devices at home to produce permanent consolidation of safety memories, these therapies can easily reach the general population and lead to new innovative approaches for promoting emotional well-being.

## Data Availability

All data produced in the present study are available upon reasonable request to the authors.

## Acknowledgements

The authors would like to thank Eleni Thomas, Ben Meuleman, Francesca Borghese, Pauline Henckaerts, Guillaume Legendre, Stephen Perrig, Virginie Sterpenich, Laurence Bayer and Kinga Igloi for useful discussions regarding this manuscript. This study was supported by the Swiss National Science Foundation, Grants/Award Numbers: CRSK-3_190722, 320030_159862, and 320030_182589.

## Author Contributions

SS and LP designed the experiments; AC and LP conducted the experiments; SS, AC and LP analyzed the data; SS, AC and LP wrote the paper.

## Materials and Methods

### Participants

Thirty-six individuals (27 women and 9 men) aged from 20 to 35 years old (M = 26, SD = 4.22) were included. Participants were recruited at the Center for Sleep Medicine, University Hospitals of Geneva. Diagnosis of ND was done by a sleep specialist according to the International Classification of Sleep Disorders (ICSD-3) diagnostic and coding manual ^3^. Patients with ND and with at least moderate severity (>1 episode per week) were included. All patients reported significant distress or impairment in social, occupational, or other important areas of functioning because of their nightmares, mainly fatigue, mood disturbance (i.e., persistence of nightmare affect) and cognitive impairments (i.e., intrusive nightmare imagery). All patients had idiopathic nightmares, as they reported that their nightmares were unrelated to a traumatic life event. A neuropsychiatric evaluation was performed to exclude patients with comorbidities, such as severe depression (measured by Beck Depression Inventory/ BDI-II ^73^), insomnia disorder, psychosis or anxiety disorder (according to DSM-5). We also excluded any patient with symptoms of obstructive sleep apnea syndrome, restless legs syndrome, neurological disease or using medications that would be likely to produce nightmares (e.g. hypnotics, β-blockers, amphetamines, antimicrobial agents). Patients with anxiolytics, antipsychotic or antidepressant medication were excluded. Signed informed consent was obtained from all participants before the experiment, and ethical approval for the study was obtained from the ethical committee of the canton of Geneva, Switzerland (*‘Commission Cantonale d’Ethique de la Recherche sur l’être humain’*).

### Procedure

The experimental design of this study is presented in Figure 1, and corresponds to a *single-blind, randomized controlled design*, with random assignment of ND patients to either the TMR group or control group (ClinicalTrials.gov identifier NCT05237778). All sessions took place at the Center for Sleep Medicine, University Hospitals of Geneva. After an initial assessment and diagnosis of ND (pre-intervention, Visit 1), all participants filled in a dream diary and a sleep diary, and wore an actigraph for two weeks. At the end of this period, all patients came to the center for a single, individual session of IRT (Visit 2). At the end of this session, participants of the TMR group received a sound (duration 1 sec, presented every 10 sec) while imagining the new positive dream scenario generated during IRT for 5 minutes. Participants from the control group also performed the 5-min imagery of the new positive dream scenario, but without sound presentation. Participants then returned home. During the following two weeks, all patients received the sound during their REM sleep. The application of sounds took place with a wearable headband device that delivered the stimulation during REM sleep (see *‘Sleep assessment’*). The patients also filled in a sleep and dream diary during these two weeks. At the end of this period, they came back to the lab for a new session during which their ND was assessed (post-intervention, Visit 3). Finally, a third assessment session took place 3 months later (3-month follow-up) (3-month follow-up, Visit 4), without any intervention between Visit 3 and 4. Twenty-eight patients (i.e., 14 subjects in each group out of the 36 participants) responded to this third assessment (dropout rate= 22%).

### Imagery Rehearsal Therapy (IRT)

A single IRT session is sufficient for the patient to learn the IRT techniques ^31^. As the main objective of the study was to assess the impact of TMR on nightmare severity, a single treatment session focusing only on nightmares (and not on psychoeducation included in the more classic 4-session IRT ^28^) was selected to avoid potential confounding effects of psychoeducation on treatment effect. Specifically, as in ^31^, information about nightmares was given to the patients and then IRT was introduced and practiced. The patients were first instructed to choose their most frequent nightmare, according their dream diary. They were then asked to change their initial nightmare in any way they wish, so that the new version would be neither unpleasant nor distressing. Then, a 5-min period was given for imagery rehearsal of the new dream. Patients were then instructed to practice imagery of the new dream scenario at home for 5 minutes per day, at least once a day, every day, during the following two weeks. During this period, patient compliance to the protocol was controlled regularly with telephone interviews.

### Group randomization, auditory stimulus and pairing procedure

The random assignment of each participant to either the TMR or the control group took place at the end of the IRT session, before the last 5 minutes of imagery rehearsal. Patients were randomly assigned to one of the two experimental groups in randomization groups of six. Sampling was done without replacement, using the “Mersenne twister” algorithm for random number generation by Matsumoto and Nishimura ^74^. The final sequence was stored in a data table and used by the investigators for the allocation to incoming participants. The sequence was concealed from the participants so that they did not know which condition they were assigned to. Participants of the TMR group received a 1-sec sound (i.e., the neutral piano chord C69, ∼40 dB) every 10 seconds through headphones while they were imagining the new positive dream scenario of IRT for 5 minutes. To avoid extinction of the sound-dream association, the patients of this group were also asked to listen to the sound every day at home while they were practicing the positive scenario of IRT. During the two weeks following Visit 2, all participants from the TMR and the control groups received the sound every 10 seconds during REM sleep (see *‘Sleep assessment’*). Before the patients returned home, and as individuals differ in their arousal threshold during sleep, the sound volume was calibrated for each participant individually (e.g., as in Sterpenich et al.^51^).

### Sleep assessment

Data from the actigraph and the sleep diary were collected from all participants for two weeks between Visit 1 and Visit 2. We could thus assess subjective sleep quality (from the diary) and sleep efficiency (TST_Acti_, SE_Acti_) (from the actigraph).

A wearable sleep headband device Dreem^®^ was then used to record sleep between Visit 2 and 3. Dreem^®^ (Dreem SAS, Paris, https://dreem.com/en/product) is a wearable sleep headband and mobile application that monitors sleep by processing the EEG signal acquired from 4 frontal and 2 occipital locations in real-time. Dreem has been previously used to apply sounds during SWS ^75^ and can reliably detect different sleep stages through an automatic algorithm ^76^. For the purpose of the present project and in collaboration with the engineers from the company, we adapted the headband to apply sounds during REM sleep. Whenever REM sleep was detected for more than five minutes, the sound was delivered to the participants every 10 seconds through bone conduction from the forehead. The sound volume during sleep was set at 10% above the individual detection threshold. As the sounds are emitted through bone conduction and not through conventional speakers, it is difficult to acquire a decibel equivalent of sound intensity. These stimulations were interrupted whenever a new sleep stage was detected (they restarted as soon as 5 minutes of stable REM were detected again), after detection of a movement (90 seconds of interruption), after detection of an alpha wave (45 seconds of interruption), after detection of a blink (10 seconds of interruption), or after bad quality signal (4 seconds of interruption). In the current protocol, the stimulation was applied at home in all participants for two weeks, between Visit 2 and Visit 3. This device also possesses an accelerometer and pulse oximeter. Its use for the two weeks of stimulation at home allowed us to calculate the number of stimulations, sound intensity, total sleep time (TST_Dreem_), sleep efficiency (SE_Dreem_), and a REM arousal index, defined here as the number of arousals in stage REM × 60/REM duration in minutes. Remote access to the raw sleep data and stimulations per night was possible, with daily controls ensuring patient compliance to the protocol.

### Dream Diary

A dream diary was given to all participants for two weeks before Visit 2 and two weeks after. This dream diary was completed every morning upon awakening and helped us detect any potential differences in emotional dream content before and after the IRT, and between groups. It contained dichotomous questions (absence/presence) for specific emotions, including fear and joy, and offered the possibility to provide an open description of the dreams (see our previous study ^22^). Based on our hypothesis that positive emotions would be reactivated by the TMR procedure, we focused our analyses on the dichotomous question about joy in the questionnaire, and its negative counterpart, fear, which may also be relevant for nightmare sufferers. We calculated (separately for each emotion) the proportion of dreams containing the emotion in the week preceding IRT (without sound stimulations) and during the week following IRT (with stimulations). To do so, we took the number of dreams where each of these emotions was present divided by the number of nights for which participants indicated that they experienced dreaming. This resulted in one proportion for each of these two emotions for the period before IRT (JOY_pre_, FEAR_pre_) and similarly for the week during IRT (JOY_post_, FEAR_post_), and this, for participants in the TMR and in the control groups.

### Questionnaires used during clinical assessments

Clinical assessment of nightmare disorder was conducted at three time points (pre-, post-, follow-up; Figure 1). In these 3 time points, we have asked the patients to retrospectively report the number of nightmares they had per week for the last 2 weeks. For this specific time frame, we have also used the validated Nightmare Distress Questionnaire (NDQ) ^77^, which evaluates the emotional disturbance attributed to the nightmares via 13 items scored on a scale ranging between “never” (0) and “always” (4). We also assessed the sleep quality and mood of our patients at the three time points via the 19-item self-rated Pittsburgh Sleep Quality Index (PSQI)^78^ and the 21-item self-rated Beck Depression Inventory II (BDI-II) ^73^, respectively. All four questionnaires have been widely used in the past to measure the efficacy of IRT ^58,59^.

### Primary and secondary outcome variables

The total number of nightmares per week was used as the primary outcome variable at post-intervention and 3-month follow-up. The proportions of joy and fear in dreams were used as secondary outcome variables (see Introduction section) at post-intervention. Other exploratory variables included the scores in NDQ, PSQI, BDI-II questionnaires, sleep efficiency (SE).

### Sample size consideration

Based on a previous study ^79^ studying the difference of nightmare patients under IRT vs patients in a control group for nightmare frequency with an effect size at d=1.01, 34 patients (17 per arm) would be required to have an 80% chance of detecting this difference between patients treated with TMR vs. those not treated. Besides, based on a study ^51^ on the effect of associated vs non-associated sound during REM sleep on associative memory with a large effect size (g=1.01), this sample size would be sufficient for the aforementioned detection.

### Statistical analyses

We first checked whether the randomization successfully generated two equivalent groups of patients in terms of their general demographic and clinical characteristics. We compared the two groups using independent t-tests for continuous variables (baseline nightmares/week, NDQ, PSQI, BDI scores, age, TST_Acti_, SE_Acti_). Independent t-tests between the two groups were also performed to check that patients from each group had similar stimulation conditions during the two weeks of stimulation at home (number of stimulations, sound intensity, SE_Dreem_, TST_Dreem_, REM arousal index, sleep stages duration). When normal distribution was not respected (Shapiro-Wilk test p<.05), Mann-Whitney U tests between the two groups were conducted.

Next, to study the effects of TMR on the outcome variables, data were entered into a multilevel regression model, with the outcome variables as dependent variables, and Time (pre-, post-, follow-up) and Group (TMR group, control group) as (interacting) independent variables. The latter represented the fixed effects of the multilevel model, while the random effects were represented by a random intercept for subjects. The random intercept accounted for correlation between repeated measures, by assuming baseline differences between subjects in the average dependent variable. A multilevel regression was chosen for these data, due to its ability to handle (a) missing data in the time variable (due to the longitudinal dropout of the current study), (b) time-varying covariates, and (c) continuous within-subject covariates.

Once the model was fitted, we performed a Type II ANOVA breakdown of fixed effects using *F*-tests, starting with the interaction test of Time × Group, followed by main effects tests for Time and Group separately. Significant effects were further explored with pairwise *t*-tests. Multiple testing correction for eventual follow-up pairwise comparisons was done using the Holm-Sidak method ^80^. Degrees of freedom for all *F*- and *t*-tests were adjusted for the random effects structure using Satterthwaite’s method ^81^, yielding fractional degrees of freedom. Multilevel analyses were conducted with the R statistical language, version 1.2.5019, (RStudio Team, Boston, MA, 2019), using the packages “lme4” for model estimation ^82^ and “lmerTest” for inferential tests ^83^.

## Supplemental information

**Figure S1**. Flow diagram of subject recruitment, randomization and analysis.

**Figure S2**. Treatment effects on the (A) NDQ score, (B) PSQI score, (C) BDI-II score and (D) proportion of fear in dreams.

**Analysis S3**. Content analysis of dreams with the Linguistic Inquiry and Word Count (LIWC)

**Analysis S4**. Latent semantic analysis (LSA) of dreams.

